# Patient characteristics associated with clinically coded long COVID: an OpenSAFELY study using electronic health records

**DOI:** 10.1101/2023.06.23.23291776

**Authors:** Yinghui Wei, Elsie MF Horne, Rochelle Knight, Genevieve Cezard, Alex Walker, Louis Fisher, Rachel Denholm, Kurt Taylor, Venexia Walker, Stephanie Riley, Dylan M Williams, Robert Willans, Simon Davy, Sebastian Bacon, Ben Goldacre, Amir Mehrkar, Spiros Denaxas, Felix Greaves, Richard J Silverwood, Aziz Sheikh, Nish Chaturvedi, Angela M Wood, John Macleod, Claire Steves, Jonathan AC Sterne, the UK COVID-19 Longitudinal Health and Wellbeing National Core Study and CONVALESCENCE study

## Abstract

Despite reports of post-COVID-19 syndromes (long COVID) are rising, clinically coded long COVID cases are incomplete in electronic health records. It is unclear how patient characteristics may be associated with clinically coded long COVID. With the approval of NHS England, we undertook a cohort study using electronic health records within the OpenSAFELY-TPP platform in England, to study patient characteristics associated with clinically coded long COVID from 29 January 2020 to 31 March 2022. We estimated age-sex adjusted hazard ratios and fully adjusted hazard ratios for coded long COVID. Patient characteristics included demographic factors, and health behavioural and clinical factors. Among 17,986,419 adults, 36,886 (0.21%) were clinically coded with long COVID. Patient characteristics associated with coded long COVID included female sex, younger age (under 60 years), obesity, living in less deprived areas, ever smoking, greater consultation frequency, and history of diagnosed asthma, mental health conditions, pre-pandemic post-viral fatigue, or psoriasis. The strength of these associations was attenuated following two-dose vaccination compared to before vaccination. The incidence of coded long COVID was higher after hospitalised than non-hospitalised COVID-19. These results should be interpreted with caution given that long COVID was likely under-recorded in electronic health records.

## INTRODUCTION

The spectrum of signs and symptoms that can newly occur or continue for months to years after severe acute respiratory syndrome coronavirus 2 (SARS-CoV-2) infection is termed long COVID^1^ or post-acute sequelae of SARS-CoV-2 (PASC)^2^. The WHO’s definition of long COVID in adults refers to signs and symptoms, usually 3 months after the onset of COVID-19, with symptoms that last for at least 2 months and cannot be explained by an alternative diagnosis^3^. In the UK context, the definition of long COVID includes both ongoing symptomatic COVID-19 (for 4-12 weeks), and post-COVID-19 syndrome (more than 12 weeks)^4^. National Institute for Health and Care Excellence guidance on supporting patients with long COVID includes assessing people with symptoms after acute SARS-CoV-2, investigations and referrals^5^.

Understanding the burden of and risk factors for long COVID is a public health priority. Counts and rates of clinically coded long COVID in English primary care varied according to demographic factors but also considerably according to the practice clinical software system^6^. This latter variation was unlikely to be explained by differences in true prevalence or case mix. Recording a long COVID code in primary care electronic health records (EHRs) can be influenced by factors including whether a patient is experiencing long COVID symptoms, their access to care, and data recording by the health workers with whom they consult.

UK longitudinal cohort studies reported that risk factors for long COVID included increasing age, female sex, obesity, poor pre-pandemic general and mental health, and asthma^7 8^. However, previous EHR analyses were based on the study period from 1 February 2020 to 9 May 2021^7^, during which 4,189 long COVID cases were clinically coded. This represents considerable under-reporting, compared with the Office for National Statistics (ONS)’s estimate of 1.0 million people with self-reported long COVID^9^ in the UK in May 2021. The usage of long COVID codes has improved with time^10^. General practice services are encouraged to enhance their knowledge on assessing and referring patients with long COVID as set out in NHS actions on long COVID for 2021/22^11^.

We conducted a cohort study within the OpenSAFELY-TPP database (https://www.opensafely.org/), which includes detailed linked data on around 24 million people registered with an English general practice (GP) using TPP SystmOne EHR software (see ‘Data source’). We aimed to quantify associations of patient characteristics, including vaccination status, COVID-19 severity, and history of a range of disease diagnoses, with coded long COVID in English primary care.

## METHODS

### Data source

We used patient data from primary care records managed by the general practice software provider, The Phoenix Partnership (TPP) SystmOne software, covering around 40% of the population in England. All data were linked, stored, and analysed securely within the OpenSAFELY platform: https://www.opensafely.org/. These data include clinically coded long COVID recorded by health and care professionals, along with information on socio-demographics, pre-existing health conditions, and frequencies of GP-patient interactions, which may be consultations or any practice contacts. Data were linked to national SARS-CoV-2 testing records (Second Generation Surveillance System), vaccination data (National Immunisation Management Service), Index of Multiple Deprivation (IMD), and the ONS death registry. Admitted Patient Care Spells (APCS) is part of Hospital Episode Statistics (HES) and is provided to OpenSAFELY via NHS Digital’s Secondary Use Service (SUS). OpenSAFELY includes pseudonymized data such as coded diagnoses, medications, and physiological parameters, but does not include free text data. Study definitions were developed in Python on GitHub, implemented in the OpenSAFELY infrastructure, and used to create a study dataset of individual patients on the OPENSAFELY secure job server (https://jobs.opensafely.org/).

### Study population and cohort definitions

Our study population consisted of all adults aged between 18 and 105 years, with known sex and region, who were registered as active patients in a TPP general practice on 29 January 2020 (the date when the first two SARS-CoV-2 cases were reported in the UK) and had at least one year of prior follow-up in a general practice, to ensure that baseline characteristics could be adequately captured.

We constructed four cohorts (Supplementary material, Figure S1, Table S1): (1) a primary general population cohort, with follow-up start date 29 January 2020 and end date the earliest of first record of any long COVID code, death date, or 31 March 2022 (the day before free SARS-CoV-2 testing in England ended^12^); (2) a post-COVID diagnosis cohort, defined regardless of vaccination status, with follow-up start date on the first recorded SARS-CoV-2 infection or COVID-19 diagnosis and end date the earliest of first record of any long COVID code, death date, or 31 March 2022; (3) a pre-vaccination cohort with follow-up start date 29 January 2020 and end date the earliest of first record of any long COVID code, date of receipt of first COVID-19 vaccine dose, death date, or 31 March 2022; (4) a post-vaccination cohort, with follow-up start date 14 days after receipt of second COVID-19 vaccine dose and end date the earliest of first record of any long COVID code, death date, or 31 March 2022.

In the primary and pre-vaccination cohorts, we followed the same population from 29 January 2020, but people in the pre-vaccination cohort were censored on the date of receipt of first COVID-19 vaccine dose. In each cohort, people with a history of SARS-CoV-2 infection and/or long COVID code prior to their follow-up start date, were excluded.

### Outcomes

The outcome was clinically coded long COVID, constructed from the date of the first record of any of the 15 UK SNOMED-CT codes for long COVID^6^ in English primary care records, consisting of two diagnostic codes, three referral codes and 10 assessment codes (Supplementary Table S2).

Assessment of long COVID was undertaken as part of routine primary care. Time to the outcome event was defined as days from participant specific follow-up start date (specified in supplementary material for each cohort in Table S1).

### COVID-19 diagnosis

Date of COVID-19 diagnosis was defined as the earliest of: record of a positive SARS-CoV-2 polymerase chain reaction or antigen test; confirmed COVID-19 diagnosis in primary care or secondary care hospital admission records; or death certificate with SARS-CoV-19 infection listed as primary or underlying cause.

### Patient characteristics

Patient characteristics included demographic variables, and health behavioural and clinical factors that may be associated with coded long COVID^6 7^, and the number of prior GP-patient interactions 12 months prior to cohort and participant specific follow-up start date, which could be an indicator of patient access to care and ability to interact with GP. There is only one entry for sex in the EHR for each patient. All other coded values were the latest recorded on or before the cohort and participant specific follow-up start date. A full description of patient characteristics is in the supplementary material, Table S2.

Demographic variables included age, sex, obesity, ethnicity, region and deprivation. Where categorised, age groups were: 18–39, 40–59, 60–79, 80–105 years. Obesity was grouped by body mass index (BMI kg m^-2^) using categories derived from the World health organization (WHO)^13^: no evidence of obesity BMI<30; obese class I, BMI 30–34.9; obese class II, BMI 35–39.9; and obese class III, BMI ≥ 40. Ethnic groups were White, Mixed, Asian or Asian British, Black or Black British and Chinese or other ethnic groups. All nine regions in England were included (East, London, East Midlands, North East, North West, West Midlands, Yorkshire and the Humber)^14^. IMD was determined based on residential area categorised into five quintiles based on relative disadvantage, with quintile 1 (Q1) being the most deprived, and quintile 5 (Q5) being the least deprived.

Health behavioural and clinical factors included smoking status, frequency of GP-patient interaction and history of disease diagnoses. Smoking status was grouped into current-, former-, and never-smokers. Frequency of GP-patient interaction was defined during the 12 months prior to participants’ follow-up start date, and categorised as: without any interaction; 1–3; 4–8; 9–12 and 13+ interactions. History of the following disease diagnoses, chosen based on previous literature on risk factors for long COVID^15^ and defined on or before the cohort and participant specific follow-up start date, was coded as separate indicator variables: asthma, cancer, chronic cardiac disease, chronic kidney disease, chronic liver disease, chronic obstructive pulmonary disease, chronic respiratory disease, dementia, diabetes, dysplenia (dysfunctional-spleen), haematological cancer, heart failure, hypertension, mental health condition, organ transplant, other immunosuppressive condition, other neurological condition, post-viral fatigue, psoriasis, rheumatoid arthritis, systematic lupus erythematosus (SLE), and stroke. History of diagnosed post-viral fatigue was defined prior to 29 January 2020 due to the potential use of the corresponding codes as a proxy for long COVID, particularly in the early stage of the pandemic prior to the introduction of specific long COVID clinical codes in December 2020.

Hospitalisation for COVID-19 was defined as a hospital admission record with confirmed COVID-19 diagnosis in primary position within 28 days of the first COVID-19 diagnosis and COVID-19 without hospitalisation as a COVID-19 diagnosis that was not followed by hospitalisation within 28 days^16^.

### Statistical analyses

Rates of coded long COVID were quantified as the number of first long COVID events per 1000 person-years. The cumulative probability of coded long COVID was estimated, using the Kaplan-Meier approach, by age group and sex. In each cohort, hazard ratios with 95% confidence intervals for each patient characteristic were estimated from age-and-sex adjusted Cox proportional hazards models, and then all patient characteristics were included in a single multivariable Cox proportional hazards model. Age was modelled using a restricted cubic spline, and estimated log hazard ratios against continuous age were plotted. In the post-COVID diagnosis cohort, we included COVID-19 severity (hospitalised vs non-hospitalised COVID-19) as an additional factor. Hazard ratios by age group (40 to 59, 60 to 79 and 80 to 105 years compared with 18 to 39 years(reference)), were estimated from models including age as a categorical variable, instead of a cubic spline.

Because of the large sample size, overfitting was expected to be minimal and so regularization of predictor effects was not considered. For computational efficiency, we used the full population with coded long COVID and a randomly sampled population without coded long COVID with an outcome-to-non-outcome ratio of 1:20. We used inverse probability weighting and robust standard errors to account for the sampling approach. The discriminative ability of the fitted model was quantified using C-statistics^17^.

We included a missing category for ethnicity, smoking status and IMD. All other covariates were defined using the presence versus absence of specific codes, and thus have no identifiable missing values.

### Data availability

All data were linked, stored, and analysed securely within the OpenSAFELY platform (https://opensafely.org/). Detailed pseudonymised patient data are potentially reidentifiable and therefore not shared. Details of access to OpenSAFELY secure data analytics platform is described on the OPENSAFELY website (https://www.opensafely.org/approved-projects/).

### Code availability

Data management and analysis were performed according to a pre-specified analysis plan, available from GitHub (https://github.com/opensafely/long-covid-risk-factors-and-prediction/tree/main/protocol) using Python 3.8 and RStudio (Professional) version 1.3 driven by R version 4.2.1. All analysis code and code lists are available from GitHub https://github.com/opensafely/long-covid-risk-factors-and-prediction). All clinical and medicines code lists are available on Open code lists (https://www.opencodelists.org/).

## RESULTS

### Study population

In total, 17,986,419 adults were included in the primary and pre-vaccination cohorts, 13,401,208 in the post-vaccination cohort and 3,507,738 in the post-COVID diagnosis cohort (<u>Table 1</u>). In the primary cohort, there were missing data for ethnicity (4,809,699, 26.74%), smoking status (744,851, 4.14%) and index of multiple deprivation (298,586, 1.66%). There were 1,855,613 (10.32%) people with ethnicity recorded as from minority groups, including Asian or Asian British, Black or Black British, Chinese or other ethnic groups, or Mixed. People in the post-vaccination and post-COVID diagnosis cohorts were more likely to have had at least one interaction with their GP 12 months prior to their follow-up than those in the primary cohort. In each cohort, the most prevalent previous diagnoses were of asthma, chronic cardiac disease, diabetes, hypertension, and mental health conditions. People in the post-vaccination cohort were older, less likely to be recorded as from a minority ethnic group, and more likely to have a history of prior disease diagnoses than those in the pre-vaccination cohort. People in the post-COVID diagnosis cohort were younger, more likely to be male, and more likely to be recorded as from a minority ethnic group than those in the primary cohort.

**Table 1.**
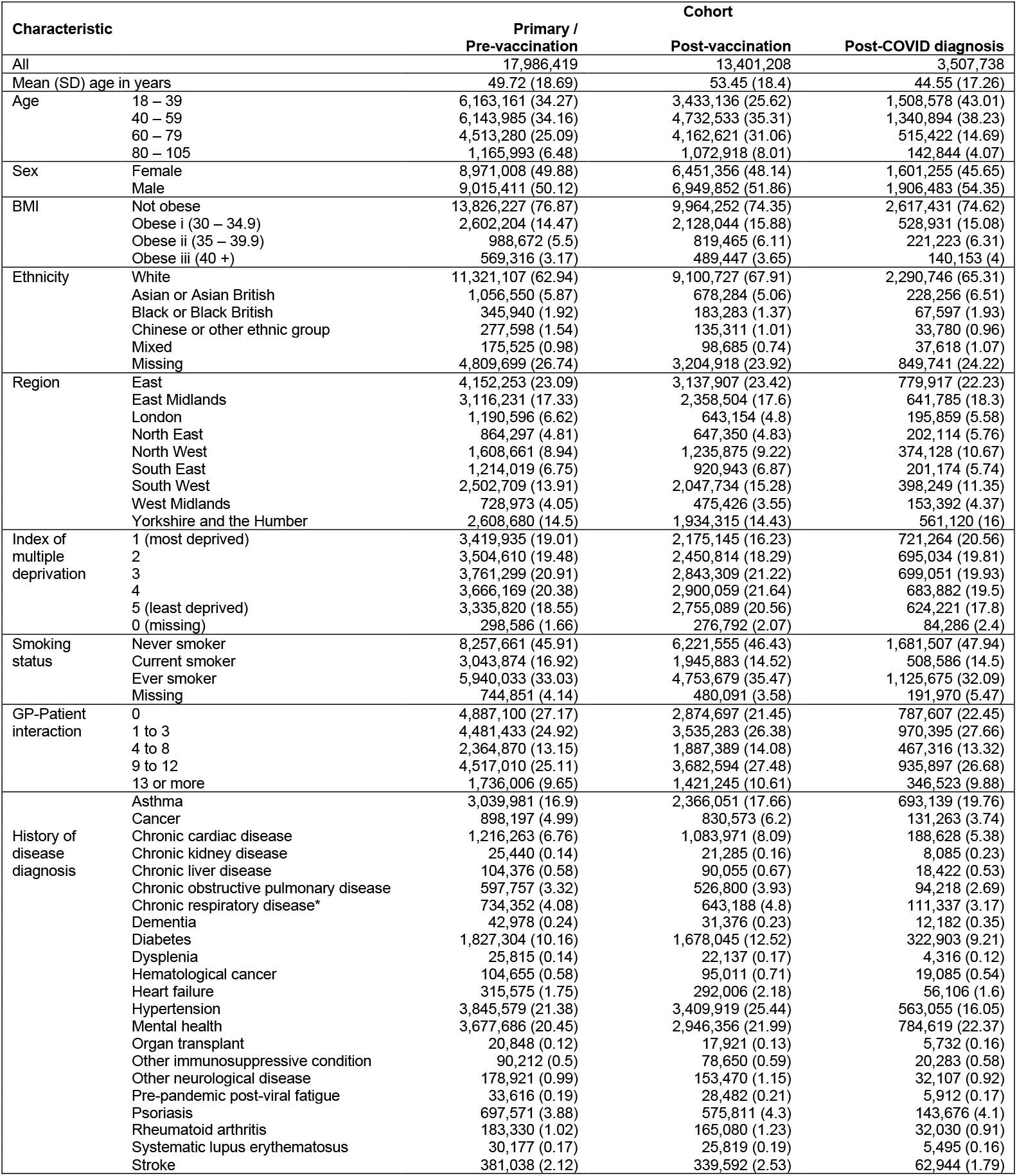
Patient characteristics. Summary statistics are number (percentage) except where indicated.

The numbers of people with coded long COVID were 36,886 (0.2%), 7,155 (0.04%), 17,376 (0.1%) and 29,268 (0.8%) in the primary, pre-vaccination, post-vaccination and post-COVID diagnosis cohorts, respectively (<u>Table 2</u>). The corresponding incidence rates of coded long COVID were 1.0, 0.3, 1.6 and 12.8 per 1000 person-years respectively. In the primary cohort, the incidence rate was highest in people aged 40-59 years (1.4), females (1.2) and people with BMI greater than 40 kg/m^2^ (1.8). In the post-COVID diagnosis cohort, the incidence rate was highest in people aged 40–59 years (17.0), females (14.8), and people with BMI greater than 40 kg/m^2^ (20.2), of white ethnicity (14.0), and living in less deprived areas (IMD Q4:14.7).

**Table 2.**
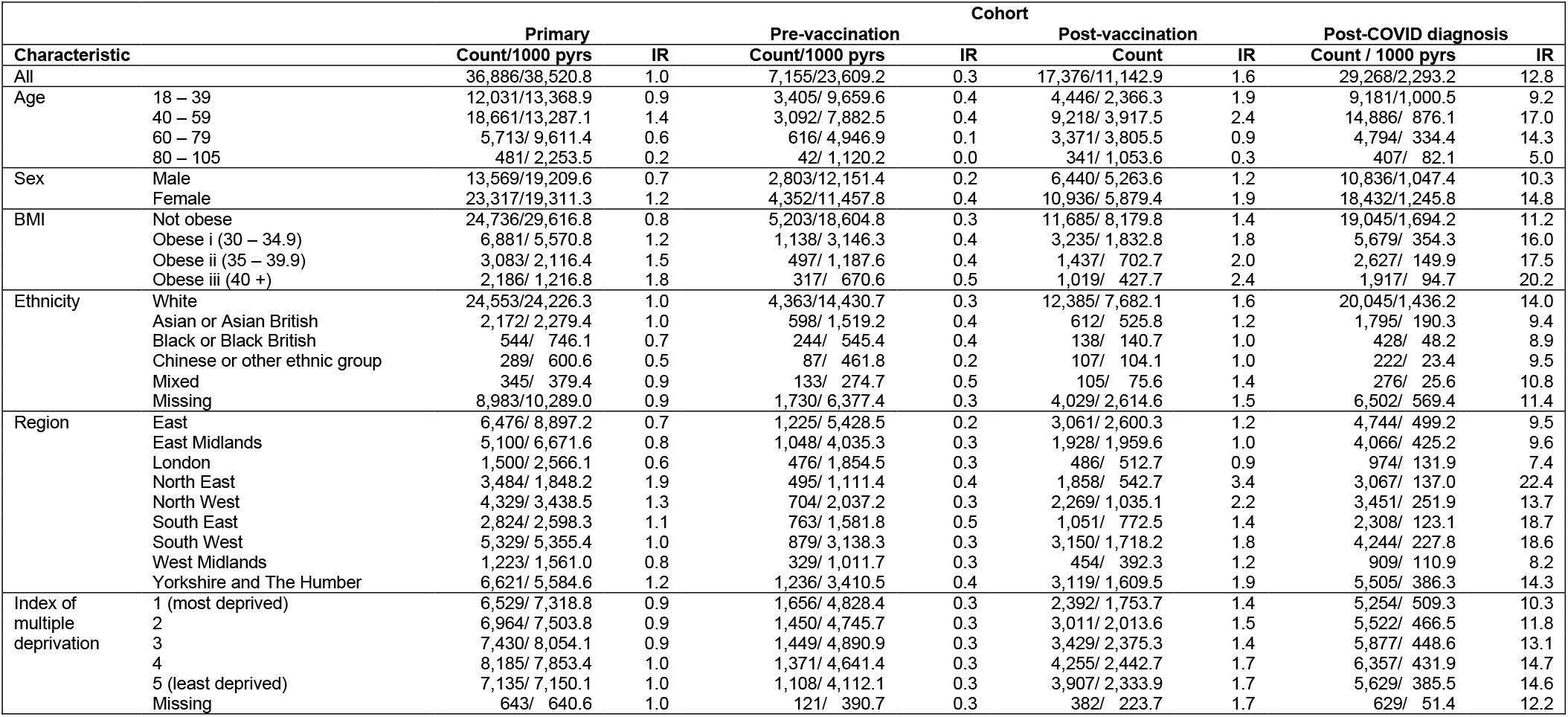
Event count /1000 person years (pyrs) and incidence rate (IR) per 1000 person years for long COVID.

In the primary cohort, the overall cumulative probability of coded long COVID was less than 0.1% in people aged 80 years or over, rising to around 0.4% and 0.2% respectively in women and men aged 40-59 years (Supplementary material, Figure S2). In the post-COVID diagnosis cohort, the overall cumulative probability of coded long COVID was less than 0.5% in people aged 80 years or over, rising to around 1.3% and 0.9% respectively in women and men aged 40-59 years (Supplementary material, Figure S3).

### Demographic factors – Primary and post-COVID diagnosis cohorts

Fully adjusted hazard ratios (aHRs) for sex, obesity and ethnicity were generally attenuated towards 1, compared with age-sex adjusted hazard ratios (Figure 1). The incidence of coded long COVID declined markedly with age in the primary cohort (aHRs 0.51 (95% CI 0.43-0.60) and 0.19 (0.15-0.24) for age groups 60-79 and 80-105 years respectively, compared with age group 18-39 years). This decline was less marked in the post-COVID diagnosis cohort. The aHRs comparing age groups were consistent with those when age was modelled by restricted cubic spline (supplementary material Figure S4). The aHRs comparing age groups were consistent with those when age was modelled by restricted cubic spline (supplementary material Figure S4). The incidence of coded long COVID was higher in females than males in (aHRs 1.33 (1.27-1.39) and 1.20 (1.14-1.27) in the primary and post-COVID diagnosis cohorts respectively). In each cohort, the incidence of coded long COVID increased with increasing obesity. In the primary cohort, the incidence of coded long COVID was lower in people from Black or Black British ethnicity (aHR 0.84 (0.74-0.96)) and Chinese or other ethnic groups (aHR 0.66 (0.56-0.77)), compared with those of white ethnicity. These differences were attenuated towards 1 in the post-COVID diagnosis cohort. In each cohort, the incidence of coded long COVID was lowest in East England, London, the East Midlands and West Midlands, and increased with decreasing deprivation.

**Figure 1.**
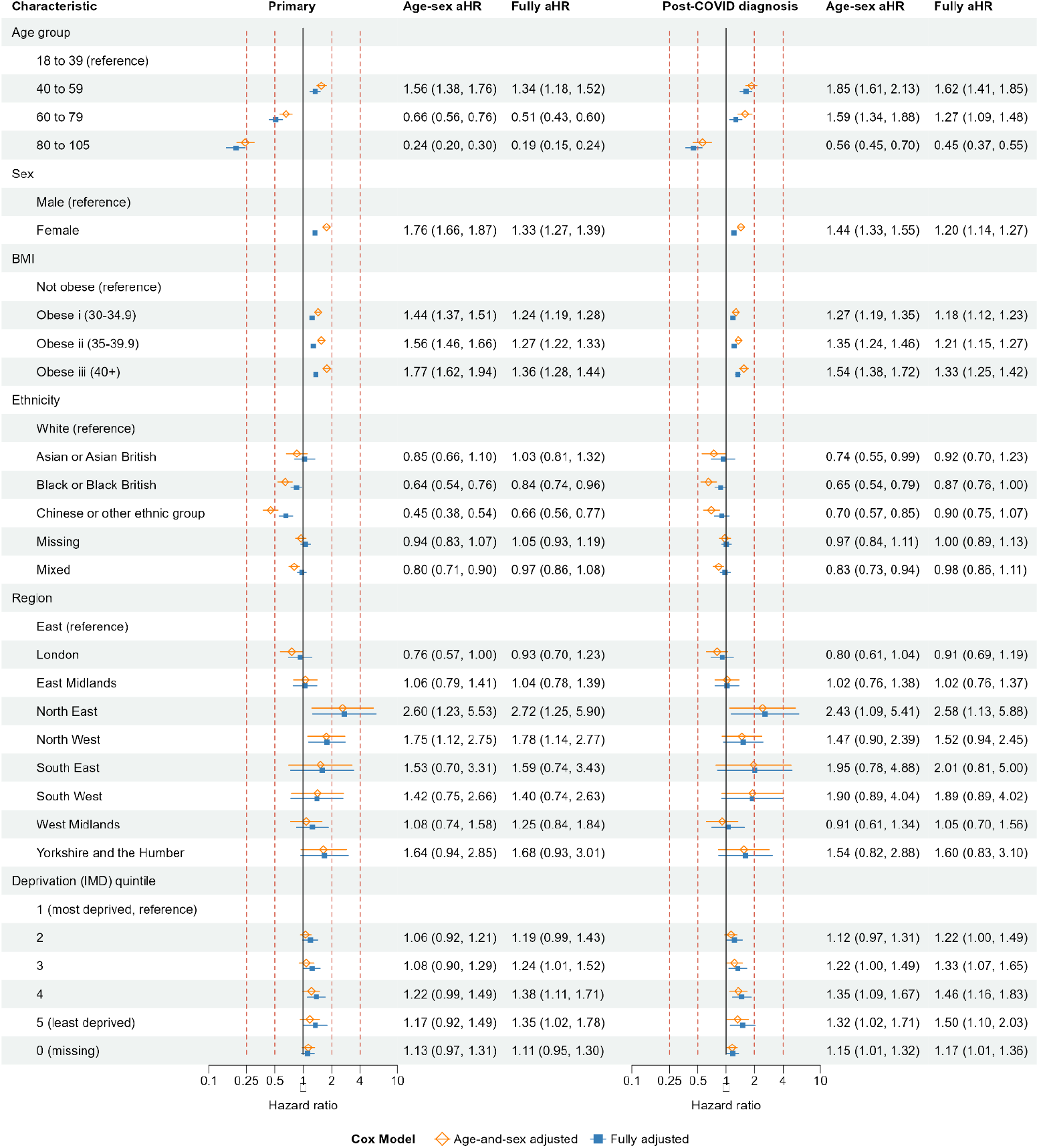
Primary and post-COVID diagnosis cohorts: age-and-sex adjusted and fully adjusted hazard ratios for demographic variables.

### Demographic factors – Pre-vaccination and post-vaccination cohorts

Fully adjusted hazard ratios (aHRs) for sex and BMI were generally attenuated towards 1, compared with age-sex adjusted hazard ratios (Figure 2). The incidence of coded long COVID declined in older adults in the post-vaccination cohort (aHRs 0.36 (95% CI 0.30-0.44) and 0.12 (0.09-0.16) for age groups 60-79 and 80-105 years respectively, compared with younger adults aged 18-39 years). This decline was less marked in the pre-vaccination cohort. The incidence of coded long COVID was higher in females than males (aHRs 1.31 (1.22-1.41) and 1.23 (1.16-1.30) in the pre-vaccination and post-vaccination cohorts respectively). In the pre-vaccination cohort, the incidence of coded long COVID was increased with increasing obesity. This pattern was less clear in the post-vaccination cohort. In both cohorts, the incidence of coded long COVID was lower in people of Chinese or other ethnic groups (aHRs 0.63 (0.50-0.81) and 0.72 (0.56-0.92) in the pre-vaccination and post-vaccination cohorts, respectively), compared with those of white ethnicity. The incidence of coded long COVID was lower in people of Black or Black British ethnicity compared to white ethnicity in the post-vaccination cohort (aHR 0.67 (0.56-0.0.81)), but not in the pre-vaccination cohort (aHR 1.10 (0.93-1.30)). In each cohort, the incidence of coded long COVID was lowest in East England, London, the East Midlands and West Midlands, and increased with decreasing deprivation.

**Figure 2.**
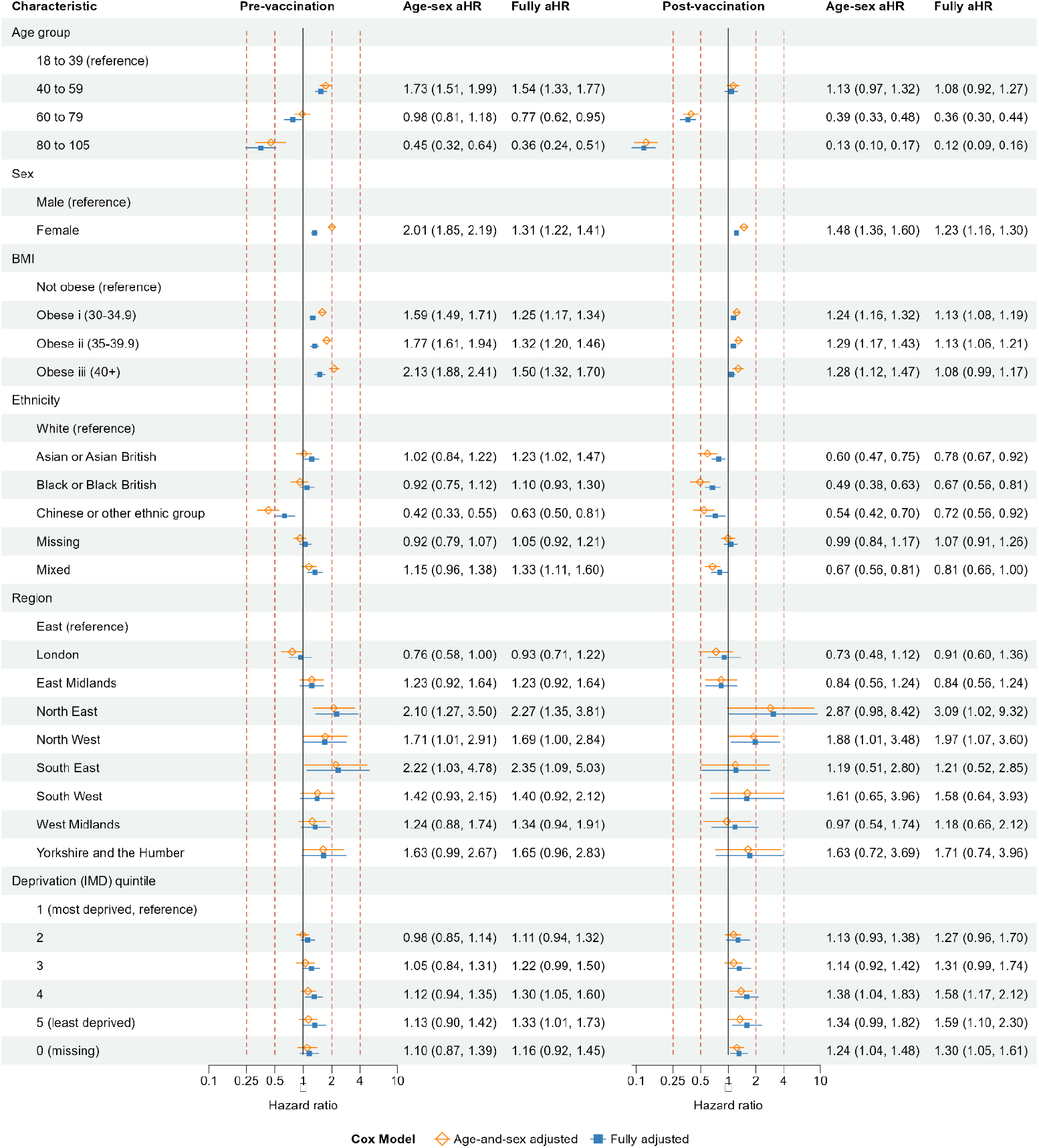
Pre-vaccination and post-vaccination cohorts: age-and-sex adjusted and fully adjusted hazard ratios for demographic variables.

### Health behavioural and clinical factors – Primary and post-COVID diagnosis cohorts

In the primary cohort, the incidence of coded long COVID was lower in current smokers and people with a missing record of smoking status, compared with people who never smoked (Figure 3). These differences were attenuated towards 1 in the post-COVID diagnosis cohort. In each cohort, the incidence of coded long COVID increased with increasing frequency of GP-patient interactions, during the 12 months prior to the follow-up start date. The aHRs for GP-patient interaction were generally attenuated, compared with age-sex adjusted hazard ratios.

**Figure 3.**
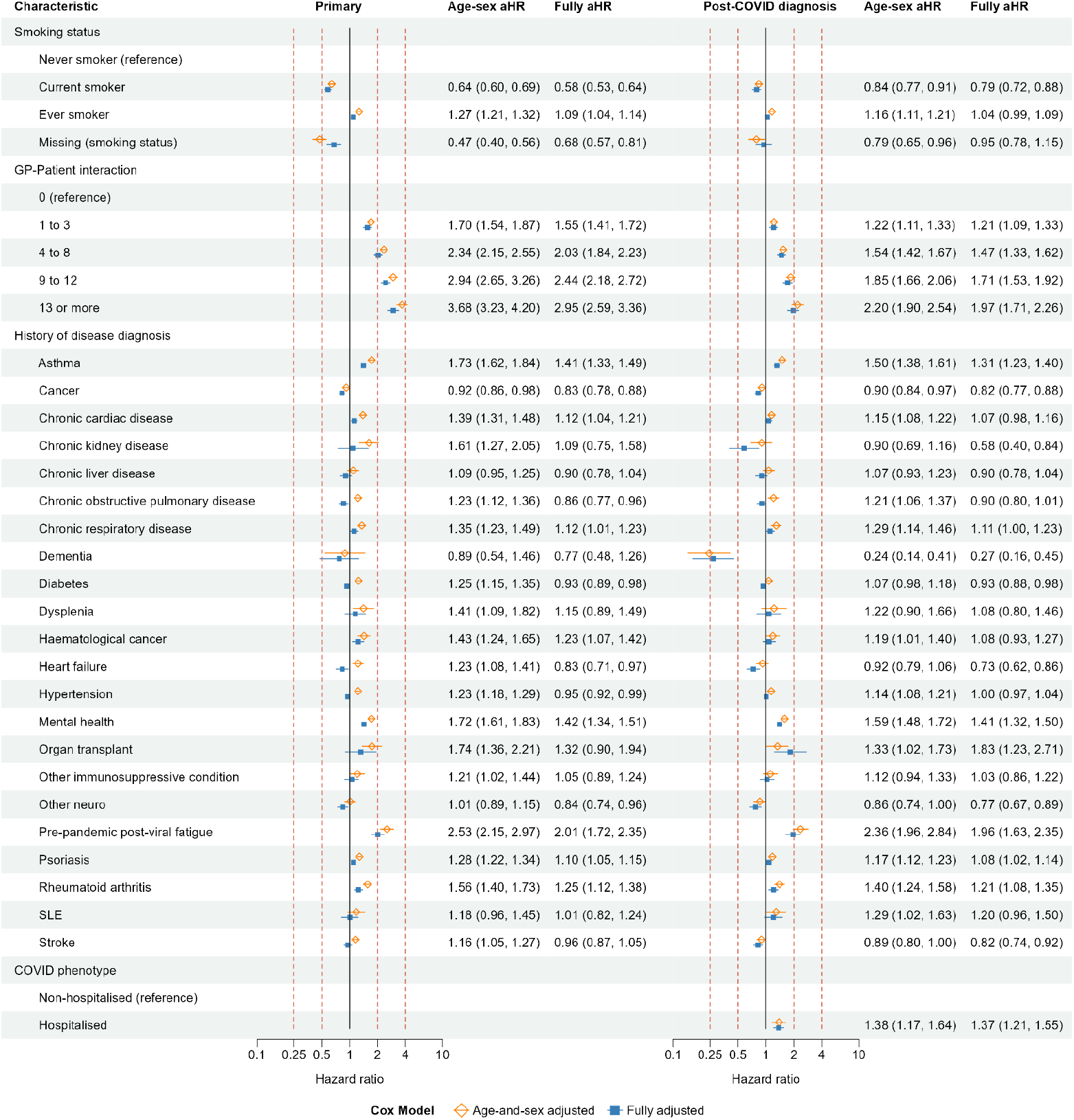
Primary and post-COVID diagnosis cohorts: age-and-sex adjusted and fully adjusted hazard ratios for health behavioural and clinical variables.

In the primary cohort, the incidence of coded long COVID was higher in people with than without a history of diagnosed asthma, chronic cardiac disease, chronic respiratory disease, haematological cancer, mental health conditions, pre-pandemic post-viral fatigue, psoriasis, or rheumatoid arthritis. These differences were generally attenuated in the post-COVID diagnosis cohort. In both cohorts, aHRs for these diseases were attenuated towards 1, compared with age-sex adjusted hazard ratios. The largest aHRs were for pre-pandemic post-viral fatigue (pre-vaccination cohort 2.01, 95% CI 1.72-2.35; post-vaccination cohort 1.96, 95% CI 1.63-2.35). In the primary cohort, the incidence of coded long COVID was lower in people with than without a history of diagnosed cancer, COPD, diabetes, heart failure, hypertension, or other neurological disorders. In the post-COVID diagnosis cohort, incidence of coded long COVID was similar in people with and without a history of diagnosed hypertension (aHR 1.00 (0.97-1.04). In the post-COVID diagnosis cohort, people who were hospitalised with COVID-19 had higher incidence of coded long COVID (aHR 1.37 (1.21-1.55)) than those who were not hospitalised.

### Health behavioural and clinical factors – Pre-vaccination and post-vaccination cohorts

In the pre-vaccination cohort, the incidence of coded long COVID was lowest in current smokers and people with a missing record of smoking status, and highest in ever smokers, compared with people who never smoked (Figure 4). The aHRs for smoking status were attenuated towards 1 in the post-vaccination cohort, compared with the pre-vaccination cohort. The incidence of coded long COVID increased with increasing frequency of GP-patient consultation, although aHRs were attenuated towards 1 in the post-vaccination cohort, compared with the pre-vaccination cohort. The aHRs for GP-patient interaction were generally attenuated, compared with age-sex adjusted hazard ratios.

**Figure 4.**
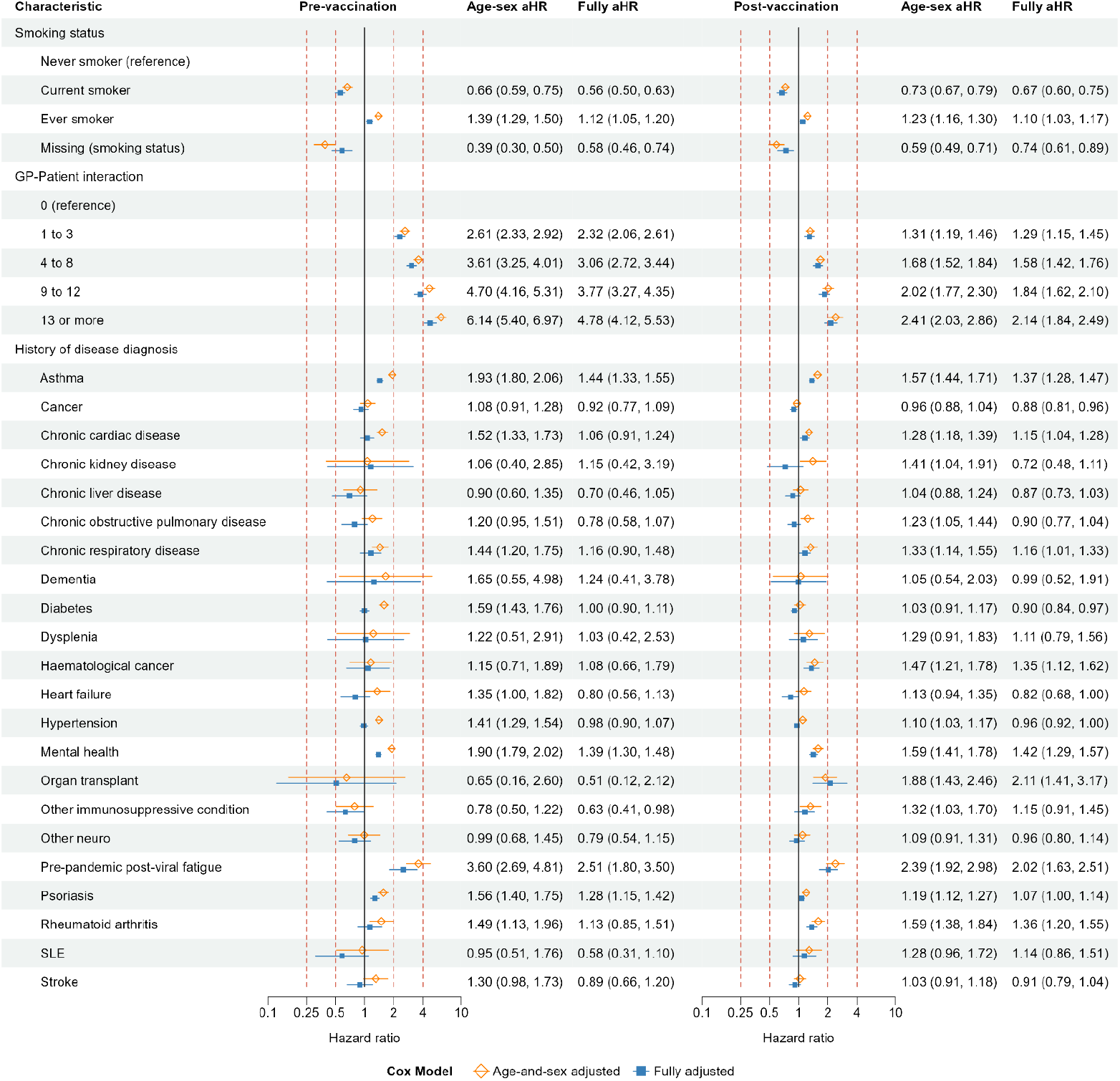
Pre-vaccination and post-vaccination cohorts: age-and-sex adjusted and fully adjusted hazard ratios for health behavioural and clinical variables.

In the pre-vaccination cohort, the incidence of coded long COVID was higher in people with than without a history of diagnosed asthma, mental health conditions, pre-pandemic post-viral fatigue, and psoriasis. These differences were attenuated in the post-vaccination cohort, compared with the pre-vaccination cohort. The aHRs for these diseases were attenuated, compared with age-sex adjusted hazard ratios. In the post-vaccination cohort, but not the pre-vaccination cohort, the incidence of coded long COVID was higher in people with than without a history of organ transplant. The incidence of coded long COVID was higher in people with than without a history of diagnosed post-viral fatigue before the pandemic, in both the pre-vaccination and post-vaccination cohorts.

## DISCUSSION

### Principal findings

Despite an estimated 2.8% of the UK population having self-reported symptoms of long COVID or post-COVID syndrome^18^ as of 3 April 2022, only 36,886 (0.2%) of the eligible general adult population in this study of up to 18 million adults had a diagnosis of long COVID recorded in their primary care record. Patient characteristics associated with higher incidence of coded long COVID included female sex, younger age (below 60 years), greater BMI, ever having smoked, and a history of diagnosed asthma, mental health conditions, and psoriasis. The incidence of coded long COVID increased with increasing frequency of prior GP-patient interactions but was lower in patients aged over 60 than under 60 years. Coded long COVID was more than twice as likely in people with than without a diagnosis of post-viral fatigue before the pandemic. The incidence of coded long COVID was higher after hospitalised than non-hospitalised COVID-19.

Differences between predictors of coded long COVID in the four cohorts studied in this paper may reflect differences between risk factors for infection with SARS-CoV-2, developing severe COVID-19, and developing long COVID having been infected with SARS-CoV-2. They may also reflect the influence of vaccination on developing long COVID, and changes in primary care coding practice and health care seeking behaviours during the pandemic. There were only minor differences between the cohorts in associations of demographic factors with coded long COVID (for example, lower incidence compared with White ethnicity for Chinese or other ethnic groups apart from the post-COVID diagnosis cohort, and for Asian or Asian British only in the post-vaccination cohort). Similarly, there were inverse associations with coded long COVID of current smoking compared with never smoking, and positive associations with number of previous GP-patient interactions, across the four cohorts, although the magnitude of this association was lower in the post-COVID diagnosis and post-vaccination cohorts than in the primary and pre-vaccination cohorts. Associations with previous disease diagnoses were also broadly consistent across the four cohorts. Taken together, these results imply that, for the risk factors studied here, the incidence of coded long COVID was mainly affected by risk factors for COVID-19, particularly severe COVID, rather than being affected by characteristics of people with COVID-19. Further, COVID-19 vaccination did not substantially modify associations of risk factors with coded long COVID-19, although it is likely to have substantially attenuated the overall incidence of COVID-19^19^.

### Strengths and limitations of this study

A key strength of this study is its use of the data from the OpenSAFELY-TPP platform, which includes over 40% of the English population^20^. We analysed data from all eligible adults in OpenSAFELY-TPP with follow-up of up to 26 months. The prevalence of coded long COVID was higher in people registered in an NHS primary care general practice using EMIS electronic health record software than in practices using TPP software^6^, which may reflect differences in the way that the software prompts structured coding. However, we were not able to access data from practices using EMIS software.

The prevalence of coded long COVID in English primary care records was substantially lower than that found in population surveys. There is likely to be considerable under-ascertainment of long COVID in these records due to difficulties in accessing care arising from health care disruption during the pandemic. Long COVID is a diagnosis of exclusion and may therefore require many interactions with health care professionals. The current lack of effective treatments may discourage patients from seeking care and primary care doctors from recording the diagnosis. Factors associated with health care access and coding of long COVID may be more influential than the risk factors for long COVID itself. Diagnoses may have been apparent in free-text despite the absence of a long COVID code, but free text was not available for our analyses. Access to free text records might help identify people with long COVID whose condition has not been coded, and thus decrease under-ascertainment^21^.

We derived and reported both age-sex adjusted and fully adjusted hazard ratios quantifying associations of demographic, as well as health behavioural and clinical characteristics with coded long COVID. Fully adjusted hazard ratios quantify the contribution of each risk factor to predicting the outcome, having accounted for the value of each other risk factors. However, they do not have causal interpretations, because they do not distinguish between adjustment for confounders (common causes of the risk factors and the outcome) and mediators (factors on the causal pathway from the risk factor to the outcome). Such misinterpretation of multiple adjusted effect estimates presented in a single table has been referred to as the ‘Table 2 Fallacy”^22^.

## Results in context with other literature

Similar to other studies^7 23 24^, we found positive associations of coded long COVID with female sex, obesity, mental health conditions and living in less deprived areas. The latter association contrasts with the increasing risk of SARS-CoV-2 infection with increasing deprivation, and illustrates the distinction between long COVID and coded long COVID, which depends on the ability of people with long COVID to access health care for their condition at a time of extreme pressure on health services. A previous EHR analysis also found that people living in less deprived areas had higher incidence of coded long COVID. However, in the same study the analysis of longitudinal cohort studies found no association between IMD and self-reported long COVID^7^.

Among the general population, the incidence for coded long COVID was lower in people of Black ethnicity, which is similar to the EHR analysis in a previous study^7^. In contrast, we found similar incidence of coded long COVID in Asian and Asian British people and people of White ethnicity. Our study additionally found that, in the general population, the incidence of coded long COVID was lower in Chinese or other ethnic groups, compared to people of White ethnicity. We found that, in general, the incidence of coded long COVID was higher in ever smokers but lower in current smokers, compared to never smokers. A previous study^7^ included only two categories for smoking status, and found no difference in the incidence of coded long COVID between current smokers and non-smokers. Smoking status in EHR may not be up to date, especially for people who had less frequent interaction with their GP.

A study in Moscow identified that pre-existing hypertension was associated with a higher risk of long COVID at 12 months follow-up since discharge from hospitalisation (OR: 1.42, 95%CI: 1.04 to 1.94)^25^. We observed that, based on the fully adjusted model in the primary cohort, the incidence of coded long COVID was lower for people with a history of diagnosed hypertension, although the incidence was higher when only adjusted for age and sex. In all other three cohorts (post-COVID diagnosis, pre-vaccination and post-vaccination cohorts), no association with hypertension was observed from the fully adjusted models. In the Moscow study, long COVID was assessed by clinicians after hospitalised COVID, whilst our study relied on people getting access to their GP and the diagnosis then being recorded.

A previous report to the UK Government’s Scientific Advisory Group for Emergencies (SAGE) also found that hospitalised COVID-19 was associated with higher risk of coded long COVID in adults, compared with non-hospitalised COVID-19^26^. Our study was restricted to adults aged 18 to 105 years. However, other studies report that hospitalised COVID was also associated with higher risks of long COVID in children^24 27 28^. A systematic review of 20 studies, which aimed to identify risk factors presented during hospitalisation for COVID, identified higher risks of long COVID with female sex, mental health conditions, fatigue and acute disease severity with respiratory symptoms^29^.

## Conclusion

Rates of coded long COVID varied by socio-demographical variable, frequency of GP-patient interaction, history of diagnosed diseases, and SARS-COV-2 severity. The results confirmed that long COVID records are incomplete in English primary care settings: under-ascertainment of long COVID poses challenges in identification of potential participants in clinical trials of interventions for long COVID. Patient characteristics associated with coded long COVID can inform evidence-based prioritisation of diagnostic assessments and clinical referrals to improve diagnosis coverage.

## AUTHOR CONTRIBUTIONS

Y.W., C.S., A.J.W. and J.A.C.S. conceptualised the study and design. Y.W., J.A.C.S., A.M.W., L.F. designed the methodology. Y.W., E.M.F.H., R.K., G.C., A.J.W. and J.A.C.S. conducted the formal analysis. Y.W., E.M.F.H., R.K., G.C., A.J.W., S.B., B.G. were responsible for data curation. Y.W., A.M. and B.G. were responsible for research ethics and information governance. S.B., S.D. and B.G. provided support for resources. S.B. and S.D. were responsible for software development. Y.W., E.M.F.H., R.K., G.C., A.J.W., R.D., K.T., V.W. and J.A.C.S. contributed to validation and visualisation. Y.W. wrote the original draft of the manuscript. All authors contributed to the review and revision of the manuscript. Project administration was conducted by Y.W., J.A.C.S., A.M., B.G., A.J.W., and L.F. The project was supervised by Y.W. and J.A.C.S. Funding was acquired by Y.W., C.S., J.A.C.S. and N.C.

## ACKNOWLEDGEMENT

We are very grateful for all the support received from the TPP Technical Operations team throughout this work, and for generous assistance from the information governance and database teams at NHS England and the NHS England Transformation Directorate. We thank the CONVALESCENCE Study Long COVID PPIE group for their input and for sharing their experiences and expertise throughout the duration of the project.

## INFORMATION GOVERNANCE AND ETHICAL APPROVAL

NHS England is the data controller for OpenSAFELY-TPP; TPP is the data processor; all study authors using OpenSAFELY have the approval of NHS England. This implementation of OpenSAFELY is hosted within the TPP environment which is accredited to the ISO 27001 information security standard and is NHS IG Toolkit compliant^30^.

Patient data has been pseudonymised for analysis and linkage using industry standard cryptographic hashing techniques; all pseudonymised datasets transmitted for linkage onto OpenSAFELY are encrypted; access to the platform is via a virtual private network (VPN) connection, restricted to a small group of researchers; the researchers hold contracts with NHS England and only access the platform to initiate database queries and statistical models; all database activity is logged; only aggregate statistical outputs leave the platform environment following best practice for anonymisation of results such as statistical disclosure control for low cell counts^31^.

The OpenSAFELY research platform adheres to the obligations of the UK General Data Protection Regulation (GDPR) and the Data Protection Act 2018. In March 2020, the Secretary of State for Health Social Care used powers under the UK Health Service (Control of Patient Information) Regulations 2002 (COPI) to require organisations to process confidential patient information for the purposes of protecting public health, providing healthcare services to the public and monitoring and managing the COVID-19 outbreak and incidents of exposure; this sets aside the requirement for patient consent^32^.

Taken together, these provide the legal bases to link patient datasets on the OpenSAFELY platform. GP practices, from which the primary care data are obtained, are required to share relevant health information to support the public health response to the pandemic and have been informed of the OpenSAFELY analytics platform.

This study was approved by NHS London - Harrow Research Ethics Committee (IRAS reference: 310808, NHS REC reference: 22/LO/0105); and by the University of Plymouth Research Ethics and Integrity Panel (reference: 3193).

