## Supplementary Material for "Patient characteristics associated with clinically coded long COVID: an OpenSAFELY study using electronic health records"

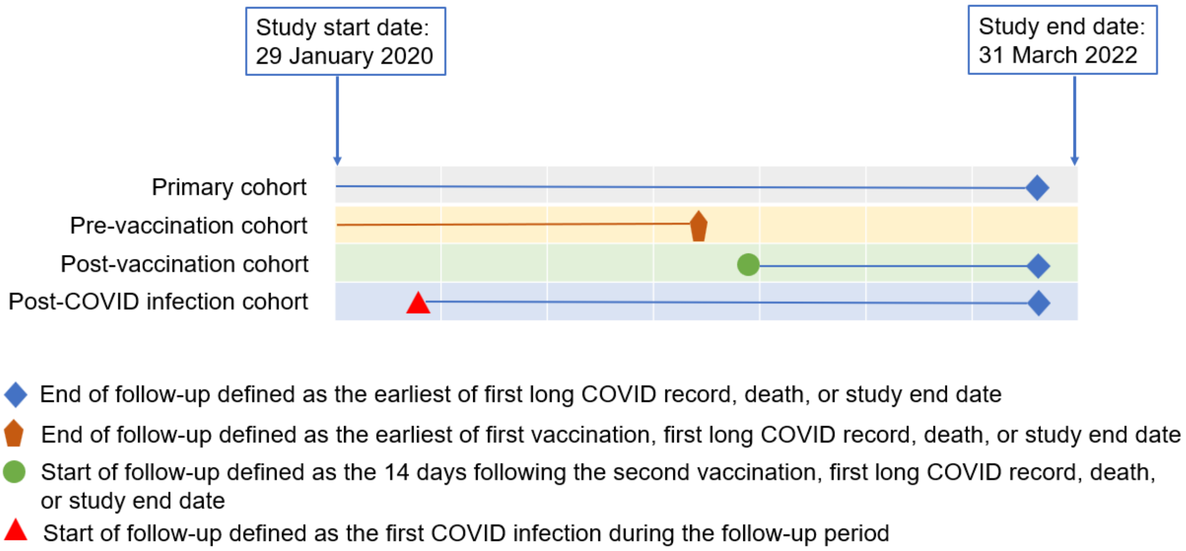


Figure S1. Illustration of follow-up start and end dates by cohort.

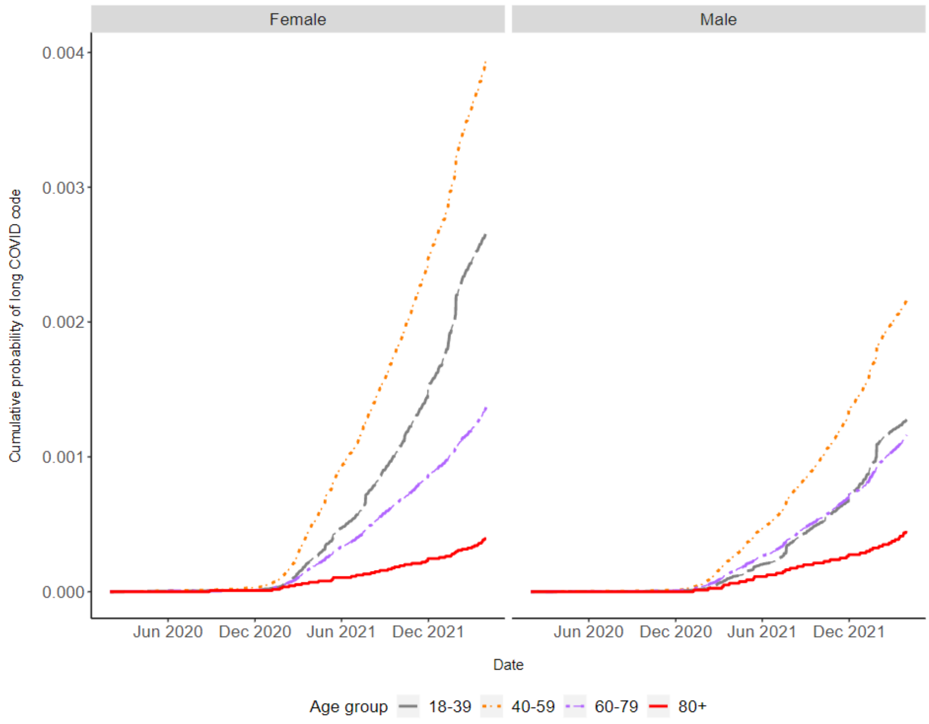


Figure S2. Kaplan-Meier plots for clinically coded long COVID for people included in the primary cohort. Plots show cumulative probability of clinically coded long COVID over time by age and sex.

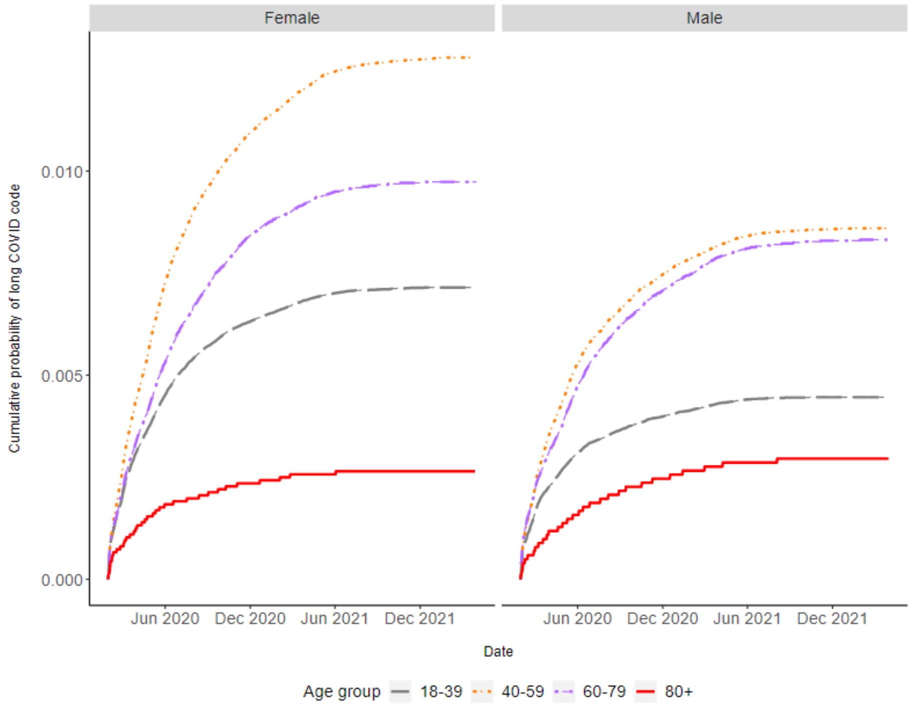


Figure S3. Kaplan-Meier plots for clinically coded long COVID for people included in the post-COVID diagnosis cohort. Plots show cumulative probability of clinically coded long COVID over time by age and sex.

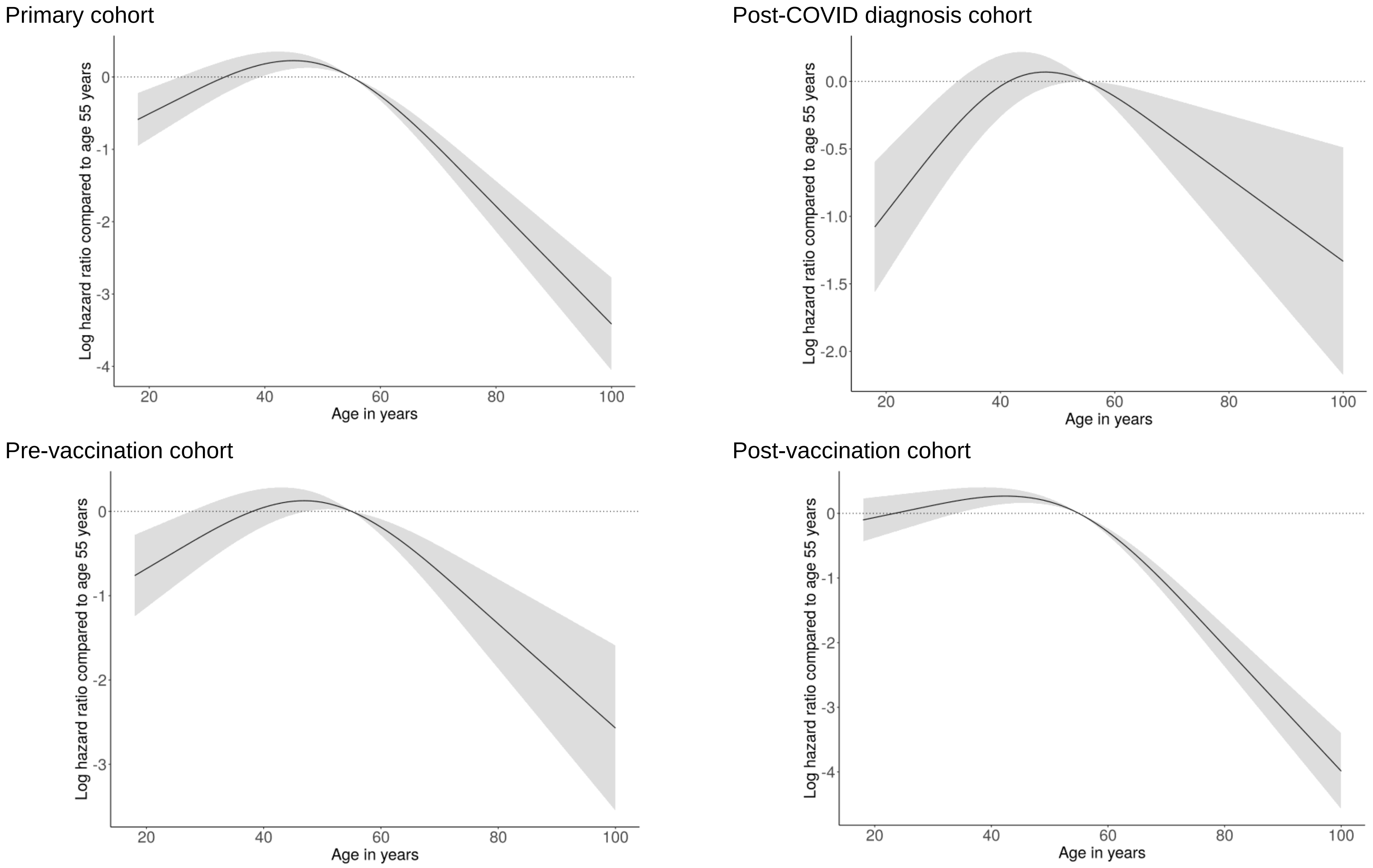


Figure S4. Estimated log hazard ratio by age in years, from Cox regression models with a three-knot cubic splines for age, and adjusted for all covariates.

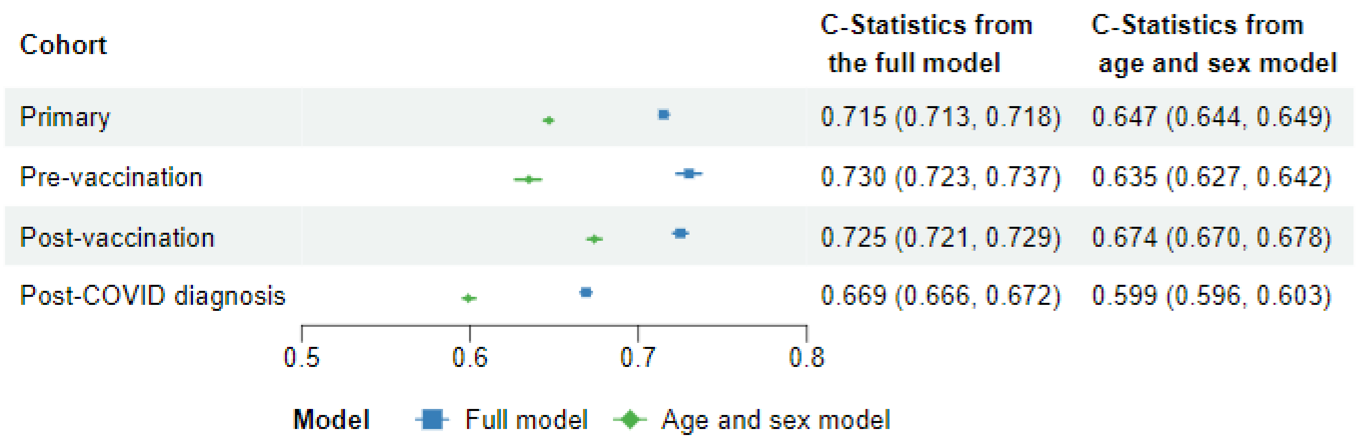


Figure S5. Apparent C statistics from Cox regression models including all covariates (full model), and including age and sex only (age and sex model) for primary cohort, pre-vaccination cohort, post-vaccination cohort and post-COVID diagnosis cohort.

Table S1. Description of study cohorts

| **Cohorts** | **Study period** | **Index date** | **Population** | **Outcome** |
|---|---|---|---|---|
| Primary cohort | 29/01/2020 to 31/03/2022 | 29/01/2020 | All eligible adults | Time from 29 January 2020 to first long COVID record, individuals censored at first long COVID record, death or end of study. |
| Pre-vaccination cohort | 29/01/2020 to 31/03/2022 | 29/01/2020 | Eligible unvaccinated adults | Time from 29 January 2020 to long COVID, individuals censored at first vaccination date, first long COVID record, death or end of study. |
| Post-vaccination cohort | 29/01/2020 to 31/03/2022 | The second vaccination + 14 days | Eligible vaccinated adults | Time from the second vaccination +14 days to first long COVID record, individuals censored at first long COVID record, death or end of study. |
| Post-COVID diagnosis cohort | 29/01/2020 to 31/03/2022 | First COVID date during the study period | All eligible adults with COVID infection | Time from COVID infection to first long COVID record, individuals censored at first long COVID record, death or end of study. |

Table S2. Long-COVID SNOMED-CT codes and terms

| **Code type and code** | **Term** |
|---|---|
| **Diagnostic codes** | |
| 1325161000000102 | Post-COVID-19 syndrome |
| 1325181000000106 | Ongoing symptomatic disease caused by severe acute respiratory syndrome coronavirus 2 |
| **Referral codes** | |
| 1325021000000106 | Signposting to Your COVID Recovery |
| 1325031000000108 | Referral to post-COVID assessment clinic |
| 1325041000000104 | Referral to Your COVID Recovery rehabilitation platform |
| **Assessment codes** | |
| 1325051000000101 | Newcastle post-COVID syndrome Follow-up Screening Questionnaire |
| 1325061000000103 | Assessment using Newcastle post-COVID syndrome Follow-up Screening Questionnaire |
| 1325071000000105 | COVID-19 Yorkshire Rehabilitation Screening tool |
| 1325081000000107 | Assessment using COVID-19 Yorkshire Rehabilitation Screening tool |
| 1325091000000109 | Post-COVID-19 Functional Status Scale patient self-report |
| 1325101000000101 | Assessment using Post-COVID-19 Functional Status Scale patient self-report |
| 1325121000000105 | Post-COVID-19 Functional Status Scale patient self-report final scale grade |
| 1325131000000107 | Post-COVID-19 Functional Status Scale structured interview final scale grade |
| 1325141000000103 | Assessment using Post-COVID-19 Functional Status Scale structured interview |
| 1325151000000100 | Post-COVID-19 Functional Status Scale structured interview |

Table S3. Description of patient characteristics.

| Variable | Type | Definition | Measurement time | Data sources |
|---|---|---|---|---|
| Age | Continuous | Age in years | Index date | Primary care |
| Age | Categorical | 18 – 39<br>40 – 59<br>60 – 79<br>80+ | Index date | Primary care |
| Sex* | Categorical | Male, Female | As per the record | Primary care |
| Obesity | Categorical | No evidence of obesity BMI<30; Obese class I, BMI 30–34.9; Obese class II, BMI 35–39.9; and Obese class III, BMI 40+. | Index date | Primary care |
| Ethnicity | Categorical | White<br>Mixed<br>Asian or Asian British<br>Black or Black British<br>Chinese or Other Ethnic Groups | Index date | Primary care |
| Region | Categorical | North East<br>North West<br>Yorkshire and the Humber<br>East Midlands<br>West Midlands<br>East<br>London<br>South East<br>South West | Index date | Primary care |
| Deprivation | Categorical | Deprivation quintiles:<br>1 (most deprived)<br>2<br>3<br>4<br>5 (least | Index date | Index of Multiple Deprivation |

| | | | | |
|---|---|---|---|---|
| | | deprived) | | |
| Smoking status | Categorical | Never smoker<br>Ever smoker<br>Current smoker | Index date | Primary care |
| GP-Patient Interaction | Categorical | Number of GP consultations in the following categories<br>0; 1-3; 4-8; 9-12; 13+ | 12 months prior to index date | Primary care |
| Asthma | Binary | Yes; No | On or before index date | Primary care |
| Cancer (exclude lung and haematological cancer) | Binary | Yes; No | On or before index date | Primary care |
| Chronic cardiac disease | Binary | Yes; No | On or before index date | Primary care |
| Chronic kidney disease | Binary | Yes; No | On or before index date | Primary care |
| Chronic liver disease | Binary | Yes; No | On or before index date | Primary care |
| Chronic obstructive pulmonary disease | Binary | Yes; No | On or before index date | Primary care |
| Chronic respiratory conditions | Binary | Yes; No | On or before index date | Primary care |
| Dementia | Binary | Yes; No | On or before index date | Primary care |
| Diabetes | Binary | Yes; No | On or before index date | Primary care |
| Dysplenia (Dysfunctional-spleen) | Binary | Yes; No | On or before index date | Primary care |
| Haematological cancer | Binary | Yes; No | On or before index date | Primary care |
| Heart failure | Binary | Yes; No | On or before index date | Primary care |
| Hypertension | Binary | Yes; No | On or before index date | Primary care |
| Mental health conditions | Binary | Yes; No | On or before index date | Primary care |
| Organ transplant | Binary | Yes; No | On or before index date | Primary care |
| Other immunosuppres | Binary | Yes; No | On or before index date | Primary care |

| | | | | |
|---|---|---|---|---|
| sive condition | | | | |
| Other neurological condition not including dementia or stroke | Binary | Yes; No | On or before index date | Primary care |
| Post-viral fatigue | Binary | Yes; No | Before pandemic start | Primary care |
| Psoriasis | Binary | Yes; No | On or before index date | Primary care |
| Rheumatoid arthritis | Binary | Yes; No | On or before index date | Primary care |
| Systemic lupus erythematosus | Binary | Yes; No | On or before index date | Primary care |
| Stroke | Binary | Yes; No | On or before index date | Primary care |

*This variable was derived once per patient without a data specification so was an exception to ‘most recent data prior to the study start date.

.
